# SARS-CoV-2 infection, risk perception, behaviour, and preventive measures at schools in Berlin, Germany, during the early post-lockdown phase: A cross-sectional study

**DOI:** 10.1101/2020.12.18.20248398

**Authors:** Franziska Hommes, Welmoed van Loon, Marlene Thielecke, Igor Abramovich, Sascha Lieber, Ralf Hammerich, Sabine Gehrke-Beck, Elisabeth Linzbach, Angela Schuster, Katja von dem Busche, Stefanie Theuring, Maximilian Gertler, Gabriela Equihua Martinez, Joachim Richter, Clara Bergmann, Alisa Bölke, Falko Böhringer, Marcus A. Mall, Alexander Rosen, Alexander Krannich, Jan Keller, Norma Bethke, Marco Kurzmann, Tobias Kurth, Valerie Kirchberger, Joachim Seybold, Frank P. Mockenhaupt, on behalf of the BECOSS study group

**Author notes:** **Corresponding author:** Franziska Hommes, Charité – Universitätsmedizin Berlin; Institute of Tropical Medicine and International Health, Augustenburger Platz 1, 13353 Berlin, Germany. Authors contributed equally (Franziska Hommes, Welmoed van Loon). The members of the BECOSS study group are acknowledged at the end of the article.

## Abstract

**Background:** Briefly before the first peak of the COVID-19 pandemic in Berlin, Germany, schools closed in mid-March 2020 for six weeks. Following re-opening, schools gradually resumed operation at a reduced level for nine weeks preceding the summer holidays.

**Aim:** During this phase, we conducted a situational assessment in schools among students and teachers as to infection status, symptoms, affective, behavioural, educational issues, and preventive measures.

**Methods:** At twenty-four randomly selected primary and secondary schools, one class each was examined. Oro-nasopharyngeal swabs and capillary blood samples were collected to assess SARS-CoV-2 infection (PCR) and specific IgG (ELISA), respectively. Medical history, household and schooling characteristics, leisure time activities, fear of infection, risk perception, hand hygiene, physical distancing, and facemask wearing were assessed.

**Results:** Among 535 participants (385 students, 150 staff), one teenager was SARS-CoV-2 infected (0.2%), and seven individuals exhibited specific IgG (1.3%); 16% reported symptoms upon examination, and 48% in the preceding 14 days. Compared to before the pandemic, the proportion of leisure time spent as screen-time increased, and the majority of primary school students reported reduced physical activity. Fear of infection and risk perception were relatively low, but acceptance of adapted health behaviours was high. Governmental preventive measures were adequately implemented, with primary schools performing better than secondary schools.

**Conclusion:** In this phase of rare infection and low seroreactivity, individual and school-level infection prevention and control measures were largely adhered to. Nevertheless, vigilance, continued and proactive preventive measures, and well-rehearsed reaction options are essential to cope with increasing pandemic activity.

## Introduction

Early in the COVID-19 pandemic, school closures were globally implemented as a central containment intervention. However, school closures bear the risk for several adverse long-term social and economic effects on children and society [1], including widening disparities [2], lowered mental health [3] and physical activity [4], and a loss of (health-care) workforce [5]. Increased screen time during school closures is observed but, so far, little is known about potentially adverse outcomes [6]. The benefits and disadvantages of school closures continue to be intensively debated [7]. In Berlin, Germany, schools attended by approximately 360,000 pupils were closed on 17 March 2020 until 27 April 2020. Following gradual re-opening, teaching under strict hygiene measures with limited pupil numbers and reduced schedules continued until 25 June, when the summer holidays started. During these nine weeks between school-reopening and summer break, SARS-CoV-2 transmission in Berlin was comparatively low; 2,523 cases were recorded by the local health authorities [8]. The reported 7-day incidence in this period varied between 2.9 and 13.9 cases/100,000 inhabitants [9].

While transmission of SARS-CoV-2 in school settings is feared, existing evidence argues against schools’ major role in actually driving rather than mirroring the pandemic. Previous data suggest that children are rarely the index cases of clusters [10–12]. In Germany, the Robert Koch Institute (i.e., the national public health institute) recorded 48 COVID-19 school outbreaks (≥2 cases) between January and August 2020, constituting 0.5% of all reported outbreaks in that period. These school outbreaks included 216 cases, of which only 30 occurred among children aged 6-10 years, whereas most cases were 21 years of age and older. As compared to the time before school closure, re-opening coincided with a slight increase of mean outbreaks per week (3.3 *vs*. 2.2) and mean cases per outbreak (6 *vs*. 4) [13]. Another analysis concluded that school re-openings had not increased the number of newly confirmed SARS-CoV-2 infections in Germany [14].

School outbreaks are recognized based on symptomatically infected individuals. However, upon SARS-CoV-2 infection, children tend to present with milder symptoms [15, 16], and up to 50% of them may stay asymptomatic [17]. The actual number of infected children attending school might thus be higher than deduced from outbreaks. On the other hand, child-to-child transmission appears to be lower than transmission from and between adults [12, 18, 19]. Limited evidence suggests that in young children (e.g., below 10 years of age) susceptibility to SARS-CoV-2 infection is lower than in adults and that infectivity increases with age. These findings are less pronounced among secondary school attendees [11]. Studies from Germany show significantly lower seroprevalence rates in young children compared with adults [17].

School-reopening in Germany was accompanied by official recommendations for infection prevention and control (IPC) measures, including health behaviours, such as hand hygiene, physical distancing, wearing face masks, or self-isolation, and testing of symptomatic students and staff. Implementation of these measures could significantly reduce disease transmission in schools [20]. Nevertheless, their effectiveness largely depends on individual adherence, which is proposed to be predicted by behaviour-related cognitions, including risk perception and fear of infection [21].

Against the background of a notable scarcity of data obtained in schools, the present study in 24 school classes in Berlin during the early post-lockdown and low incidence phase aimed at assessing parameters describing the changed schooling and living situation. These parameters included current and recent signs and symptoms, the prevalence of SARS-CoV-2 infection and seroprevalence, household composition and leisure activities, risk perception as to SARS-CoV-2 infection, adherence to infection prevention health behaviours, and school-level infection prevention measures.

## Methods

### Study design, setting and participants

This was a cross-sectional study performed in 24 Berlin schools between June 11 and 19, 2020. For the selection of schools, the twelve districts of Berlin were divided into three socio-economic strata according to the city’s Social Atlas [22]. In a random selection process, two districts *per* stratum and in each selected district, two primary and two secondary schools were chosen. Three schools refused to participate (two because of organisational concerns, one because of an expected low participation rate) and were replaced by substitutes of the same stratum. *Per* facility, one class was chosen by the school to account for organisational necessities. In primary schools, classes were selected amongst grades 3-5, in secondary schools amongst grades 9-11. We aimed at examining 20 students *per* class and up to 10 school-staff with direct contact to the class (teachers, educators). Written study information was provided to potential participants at least one week prior to the school visit to obtain written consent from parents or legal guardians, and study staff was available for questions *via* telephone.

### Data collection

Study teams visited the schools at scheduled dates. At the visits, body temperature was measured by forehead scanners, with a temperature of ≥37.5 degrees Celsius defined as fever. A brief medical history was obtained, including fever, acute respiratory symptoms, and loss of smell or taste. Combined oropharyngeal/naso-pharyngeal swabs (eSwab, Copan, Italy) were professionally collected, and finger-prick blood samples were taken onto filter-paper (Bio Sample Card, Ahlstrom Munksjö, Germany). Consenting study participants who were absent during the school visit due to reported disease were visited at home, usually on the same day. SARS-CoV-2 infection was determined by real-time-PCR (Roche Diagnostics, Switzerland) within 24 hours after swab collection. Anti-SARS-CoV-2-IgG was assessed by punching 4.75 mm discs from dried blood spots, extracting samples in 250 µl sample buffer at ambient temperature for 1 hour, and ELISA was performed on a EUROLabWorkstation (Euroimmun AG, Germany).

A week before the study visit, participants were asked to complete a paper-based questionnaire (adapted versions for children, adolescents, and adults) assessing, among other variables, signs and symptoms, household composition, contacts to positive cases, risk factors, fear of infection, risk perception towards SARS-CoV-2 infection, health behaviours including hand hygiene, physical distancing and facemask wearing, and leisure time activities, the latter item comparing the present to the pre-pandemic situation. One-item assessments were used; response scales in aggregated form are depicted in Tables 1 and 2.

**Table 1.** Reported symptoms on examination day, during the preceding 14 days, and medical history.

|  | Primary school students (N=193) |  | Secondary school students (N=192) |  | Staff (N=150) |  | Total (N=535) |  |
| --- | --- | --- | --- | --- | --- | --- | --- | --- |
| On examination day: | % | n/N | % | n/N | % | n/N | % | n/N |
| Any of the defined symptoms | 18.8 | 36/192 | 16.3 | 31/190 | 12.2 | 18/148 | 16.0 | 85/530 |
| Headache | 6.8 | 13/192 | 6.8 | 13/192 | 7.4 | 11/148 | 7.0 | 37/532 |
| Rhinorrhoea | 7.8 | 15/192 | 7.8 | 15/192 | 2.0 | 3/148 | 6.2 | 33/532 |
| Cough | 5.7 | 11/192 | 2.6 | 5/192 | 3.4 | 5/148 | 3.9 | 21/532 |
| Sore throat | 3.1 | 6/192 | 3.6 | 7/192 | 0.7 | 1/148 | 2.6 | 14/532 |
| Diarrhoea | 2.1 | 4/192 | 0 | 0/192 | 1.4 | 2/148 | 1.1 | 6/532 |
| Limb pain | 1.0 | 2/192 | 0 | 0/192 | 0 | 0/148 | 0.4 | 2/532 |
| Loss of smell and taste | 0.5 | 1/190 | 0.5 | 1/190 | 0 | 0/148 | 0.4 | 2/528 |
| Self-reported fever | 1.6 | 3/192 | 0 | 0/192 | 2.0 | 3/148 | 1.1 | 6/532 |
| Fever $\geq 37.5^{\circ}\text{C}$; measured | 2.1 | 4/192 | 3.1 | 6/192 | 1.3 | 2/149 | 2.3 | 12/533 |
| **During preceding 14 days:** |  |  |  |  |  |  |  |  |
| Any of the defined symptoms | 40.6 | 76/187 | 54.9 | 101/184 | 48.0 | 71/148 | 47.8 | 248/519 |
| Headache | 21.5 | 41/191 | 35.4 | 67/189 | 32.9 | 49/149 | 29.7 | 157/529 |
| Rhinorrhoea | 11.1 | 21/190 | 20.2 | 38/188 | 7.5 | 11/146 | 13.4 | 70/524 |
| Cough | 5.3 | 10/190 | 11.6 | 22/189 | 4.7 | 7/149 | 7.4 | 39/528 |
| Sore throat | 7.0 | 13/187 | 14.4 | 27/187 | 6.9 | 10/145 | 9.6 | 50/519 |
| Diarrhoea | 7.0 | 13/186 | 4.9 | 9/184 | 7.6 | 11/145 | 6.4 | 33/515 |
| Limb pain | 3.7 | 7/189 | 5.4 | 10/186 | 3.5 | 5/144 | 4.2 | 22/519 |
| Loss of smell and taste | 0 | 0/191 | 1.6 | 3/185 | 0 | 0/147 | 0.6 | 3/523 |
| Breathlessness | 4.7 | 9/192 | 11.1 | 21/189 | 6.1 | 9/148 | 7.4 | 39/529 |
| Tiredness | 4.8 | 9/188 | 8.0 | 15/188 | 18.8 | 28/149 | 9.9 | 52/525 |
| Chills | 0 | 0/187 | 1.6 | 3/187 | 2.1 | 3/146 | 1.2 | 6/520 |
| Chest pain | 1.6 | 3/188 | 4.3 | 8/187 | 3.4 | 5/146 | 3.1 | 16/521 |
| Self-reported fever | 0.5 | 1/190 | 0.5 | 1/189 | 2.0 | 3/149 | 0.9 | 5/528 |
| **Self-reported chronic diseases** |  |  |  |  |  |  |  |  |
| Any | 13.0 | 25/192 | 18.1 | 34/188 | 44.2 | 65/147 | 23.5 | 124/527 |
| High blood pressure | 0 | 0/193 | 0 | 0/192 | 14.0 | 21/150 | 3.9 | 21/535 |
| Chronic heart disease | 0 | 0/193 | 2.1 | 4/192 | 4.7 | 7/150 | 2.1 | 11/535 |
| Obesity | 1.0 | 2/193 | 0 | 0/192 | 6.7 | 10/150 | 2.2 | 12/535 |
| Diabetes mellitus | 0 | 0/193 | 0 | 0/192 | 0.7 | 1/150 | 0.2 | 1/535 |
| Chronic lung disease | 4.1 | 8/193 | 4.2 | 8/192 | 4.0 | 6/150 | 4.1 | 22/535 |
| Immunodeficiency | 0.5 | 1/193 | 0.5 | 1/192 | 1.3 | 2/150 | 0.7 | 4/535 |
| Cancer | 0 | 0/193 | 0 | 0/192 | 2.0 | 3/150 | 0.6 | 3/535 |
| Other | 8.8 | 17/193 | 10.4 | 20/192 | 21.3 | 32/150 | 12.9 | 69/535 |
| Regular medication | 7.8 | 15/192 | 12.2 | 23/188 | 39.3 | 57/145 | 18.1 | 95/525 |
| Allergies | 29.4 | 55/187 | 26.5 | 48/181 | 40.7 | 57/140 | 31.5 | 160/508 |
| Smoking | - |  | 5.3 | 10/188 | 23.8 | 35/147 | 13.4 | 45/335 |

**Table 2.** Proportions for reported fear of infection, risk perception, and health behaviours.

|  | **Students**<br>**primary school**<br>**(N=193)** |  | **Students**<br>**secondary**<br>**school**<br>**(N=192)** |  | **Staff**<br>**(N=150)** |  | **Total**<br>**(N=535)** |  |
| --- | --- | --- | --- | --- | --- | --- | --- | --- |
| **Fear of infection** | **%** | **n/N** | **%** | **n/N** | **%** | **n/N** | **%** | **n/N** |
| Not at all to a bit | 72.9 | 137/188 | 70.0 | 133/190 | 51.7 | 77/149 | 65.8 | 347/527 |
| Medium to very strong | 27.1 | 51/188 | 30.0 | 57/190 | 48.3 | 72/149 | 34.2 | 180/527 |
| **Risk perception** |  |  |  |  |  |  |  |  |
| Very low to rather low | 72.9 | 137/188 | 60.5 | 115/190 | 40.9 | 61/149 | 59.4 | 313/527 |
| Medium to very high | 27.1 | 51/188 | 39.5 | 75/190 | 59.1 | 88/149 | 40.6 | 214/527 |
| **Hand washing/using handrub per day** |  |  |  |  |  |  |  |  |
| 0-1 time | 1.0 | 2/192 | 4.7 | 9/191 | 0 | 0/149 | 2.1 | 11/532 |
| 2-4 times | 34.4 | 66/192 | 39.3 | 75/191 | 12.1 | 18/149 | 29.9 | 159/532 |
| ≥ 5 times | 64.6 | 124/192 | 56.0 | 107/191 | 87.9 | 131/149 | 68.0 | 362/532 |
| **Physical distancing at school** |  |  |  |  |  |  |  |  |
| Never to rarely | 2.6 | 5/189 | 15.8 | 30/190 | 3.4 | 5/147 | 7.6 | 40/526 |
| Sometimes | 16.4 | 31/189 | 29.5 | 56/190 | 19.7 | 29/147 | 22.1 | 116/526 |
| Frequently to always | 81.0 | 153/189 | 54.7 | 104/190 | 76.9 | 113/147 | 70.3 | 370/526 |
| **Facemask wearing at school** |  |  |  |  |  |  |  |  |
| Never to rarely | 38.9 | 74/190 | 53.7 | 102/190 | 38.5 | 57/148 | 44.1 | 233/528 |
| Sometimes | 11.1 | 21/190 | 11.1 | 21/190 | 21.6 | 32/148 | 14.0 | 74/528 |
| Frequently to always | 50.0 | 95/190 | 35.3 | 67/190 | 39.9 | 59/148 | 41.9 | 221/528 |
| **Situations of facemask wearing in the school context multiple response scale)** |  |  |  |  |  |  |  |  |
| Never | 32.9 | 55/167 | 29.4 | 55/187 | 25.5 | 36/141 | 29.5 | 146/495 |
| In class | 38.3 | 64/167 | 9.6 | 18/187 | 54.6 | 77/141 | 32.1 | 159/495 |
| In schoolyard | 32.9 | 55/167 | 35.8 | 67/187 | 22.7 | 32/141 | 31.1 | 154/495 |
| On way to school | 24.0 | 40/167 | 48.1 | 90/187 | 24.8 | 35/141 | 33.3 | 165/495 |

Lastly, school-level IPC measures were documented to examine the grade of implementation of official recommendations [23]. For that, class teachers completed a questionnaire on implementation of these IPC measures, including 1) basic hygiene measures, including hand hygiene; 2) keeping distance; 3) absence rule for symptomatic persons; 4) fresh-air-ventilation, at least once/school break; 4) cohort building of learning groups; 5) changes to the subjects taught: physical education outside, no choir/theatre/orchestra rehearsals; 6) staggering of teaching hours; 7) home-schooling for staff and students belonging to risk groups.

### Data processing and statistical analysis

All data collection was done in a pseudonymised manner on paper forms, subsequently digitalised and managed using REDCap electronic data capture tools hosted at Charité – Universitätsmedizin Berlin [24]. Descriptive analyses were segregated for primary school students, secondary school students, and school staff.

### Ethical statement

This study was reviewed and approved by the ethics committee of Charité – Universitätsmedizin Berlin (EA2/091/20) and informed written consent and assent was obtained from all participants and their legal guardians in the case of minors.

## Results

### Participants

In 12 primary and 12 secondary schools, 535 participants were enrolled in the study, including 36% (193/535) primary school students, 36% (192/535) secondary school students, and 28% (150/535) school-staff. The inclusion rate of students (i.e., enrolled students by all students *per* class) was 65% (range, 13%-96%). Median age (range) was 10 (8-13) years for primary school students and 15 (13-18) for secondary school students; half (190/383) were male. Staff participants comprised of 76% (112/148) teachers and educators and 24% (36/148) facilitating personnel. The majority of school staff was female (71%; 98/138), and aged 41-50 years (29%; 43/150). Compliance with sample collection was high: none of the staff refused the swab and only 0.7% (1/150) the finger-prick; among students, 0.8% (3/385) refused the swab and 0.8% (3/385) the finger-prick.

### Prevalence of SARS-CoV-2 infection and IgG antibodies

We detected one SARS-CoV-2 infection among 532 participants (0.2%): a 16-year old, afebrile female student who reported headache and rhinorrhoea as only symptoms upon enrolment. For the 14 days before enrolment, she reported cough, headache, rhinorrhoea and limb pain, and she was unaware of any contact to a positive case. Seven participants (1.3%; 7/527), all students, showed IgG antibodies to SARS-CoV-2, three of them belonged to one secondary school class. The median (range) age of the sero-reactive students was 14 (9-17) years and 5/7 were female. One of them reported loss of smell and taste within the preceding two weeks.

### Signs and symptoms, and chronic conditions

At examination, fever was present in 2.3% of all participants. Any sign or symptom on the examination day was reported by 19% of primary school students, 16% of secondary school students, and 12% of school staff (Table 1). Leading symptoms were headache, rhinorrhoea, cough, and sore throat. Individual symptoms differed between students and school staff, and loss of smell or taste was reported rarely and exclusively by students (0.5%) (Table 1). Signs and symptoms in the preceding 14 days were reported by nearly half of the participants (48%), with headache, rhinorrhoea and sore throat leading and with differences between the three groups of participants (Table 1).

One in four individuals (24%) reported having a chronic disease or condition, substantially more so among staff (44%). Among those with a chronic condition, 18% had a chronic lung disease, 17% high blood pressure, 10% stated to be obese, 9% had chronic heart disease, 3% an immune disease, 2% cancer, and 0.8% diabetes mellitus (Table 1).

### Household characteristics and leisure activities

Most participants lived in households of 3-4 persons. Households rather comprised kindergarten children (19.0%, 94/495) than people above the age of 60 years (13.1%, 64/489). Most students (79.4%, 305/384) had a room for themselves. Contacts to a confirmed (0.8%, 4/527) or suspected (1.9%, 10/520) COVID-19 case in the preceding 14 days were rarely reported. The main way of transport to school was walking for primary school students (53.2%, 101/190), public transport for secondary students (50.5%, 96/190), and car driving for staff (54% (81/150).

Changes in leisure time activities compared to pre-pandemic times are displayed in Figure 1. Spending time with friends was greatly reduced across all three subgroups (63%-81%). A clear reduction of physical activity was seen for primary school students only. In contrast, substantial proportions of students reported increases in the use of social media and “screen-time” (e.g., gaming and watching TV). For instance, more than 40% of the students stated spending more time with YouTube and TV than previously. Screen-time increased particularly among children compared to adults (Figure 1).

**Figure 1.**
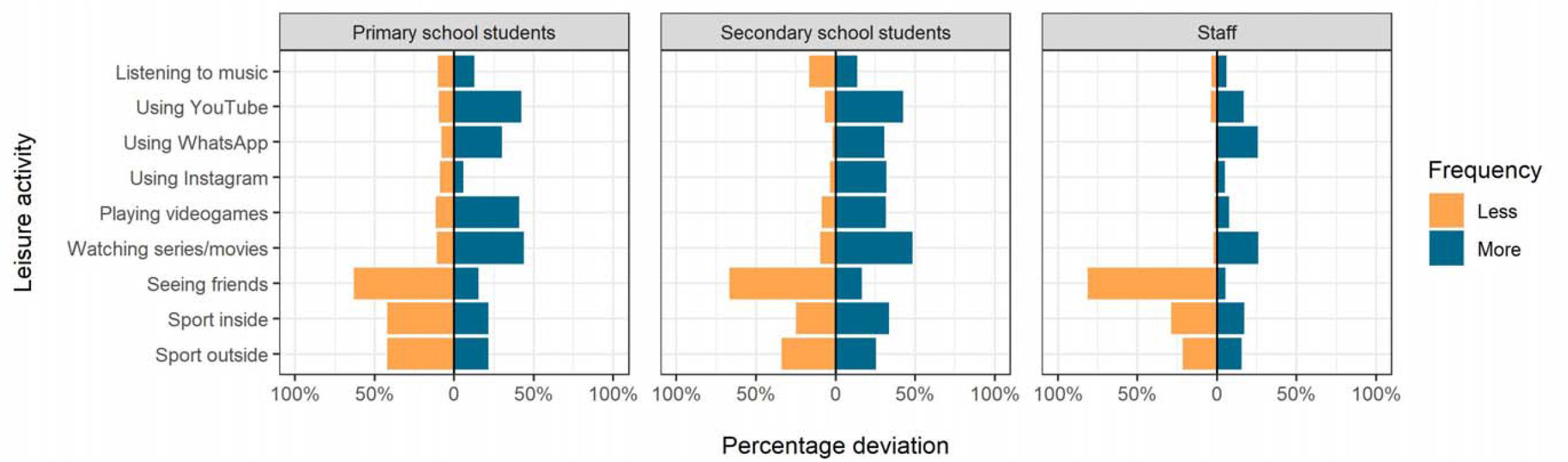
Deviations of time spent on leisure activities within preceding 14 days as compared to pre-pandemic times. **Note**. Missing values to 100% reflect no change of activities.

### Fear of infection and risk perception

Two-thirds of all participants expressed no or only little fear of infection, and a similar proportion perceived no or a rather low personal risk to contract SARS-CoV-2 (Table 2). Of note, fear and risk perception were pronounced among staff: half of them reported moderate to very high fear of infection and 59% reported moderate to very high perceived risk of infection, whereas these figures were lower among students. Moreover, fear of infection was significantly correlated with increased age and the presence of chronic conditions among staff (two-tailed Cochran-Armitage test, Z-statistic, -2.2; p=0.03; and Fisher’s exact test, OR, 2.5; 95% CI 1.2-5.1).

### Individual-level health behaviours

Overall, recommended individual-level health behaviours were adhered to well (Table 2): 68% of all participants reported to wash their hands or use disinfection handrub at least 5 times per day. Among school staff, 88% fell in this response range, whereas 65% of primary and 56% of secondary school students did. Physical distancing at school and in public was followed frequently or always by over 70% of all participants. As with handrub use, this proportion was highest among staff, less among primary school students, and least in older students (Table 2). The highest proportion of frequent or continuous facemask use in school was found among primary school students (50%), followed by older students (35%) and school staff (40%). About one third of the total sample (34%) reported never wearing facemask at school. Of all participants, 72.3% (349/483) reported wearing fabric or cloth masks followed by 41% (198/483) reporting to wear surgical masks (on a multiple response scale).

### School-level infection prevention measures

Data on the implementation of obligatory IPC measures and recommendations in the visited schools are displayed in Figure 2. Highest adherence rates were observed for keeping distance and fresh air ventilation. Basic hygiene measures, such as daily cleaning of the classroom, were implemented at every school, but less than half of schools had a hygiene commissioner. While the majority of schools had reduced class sizes at the time of the study, class cohorting outside of the classroom was practiced in less than half of the facilities. In primary schools, sports activities were suspended, and only a minority of secondary school classes had physical education outside instead of inside. All schools implemented measures going beyond governmental requirements at that time including wearing facemask, teaching hours outdoors, restricted parental access, daily documentation of absent staff and students, and closure of the canteen. The implementation rate of these additional measures was very heterogeneous. Overall, more primary schools implemented preventive measures compared to secondary schools (Figure 2). As for distance learning, 67% (14/21) of the classes reported some kind of online teaching. On average, 15% (range, 0-50%) of teaching was held online at primary schools and 50% (range, 0-90%) at secondary schools. In total, three classes (13%; all at primary schools) reported that all persons in the class were wearing masks.

**Figure 2.**
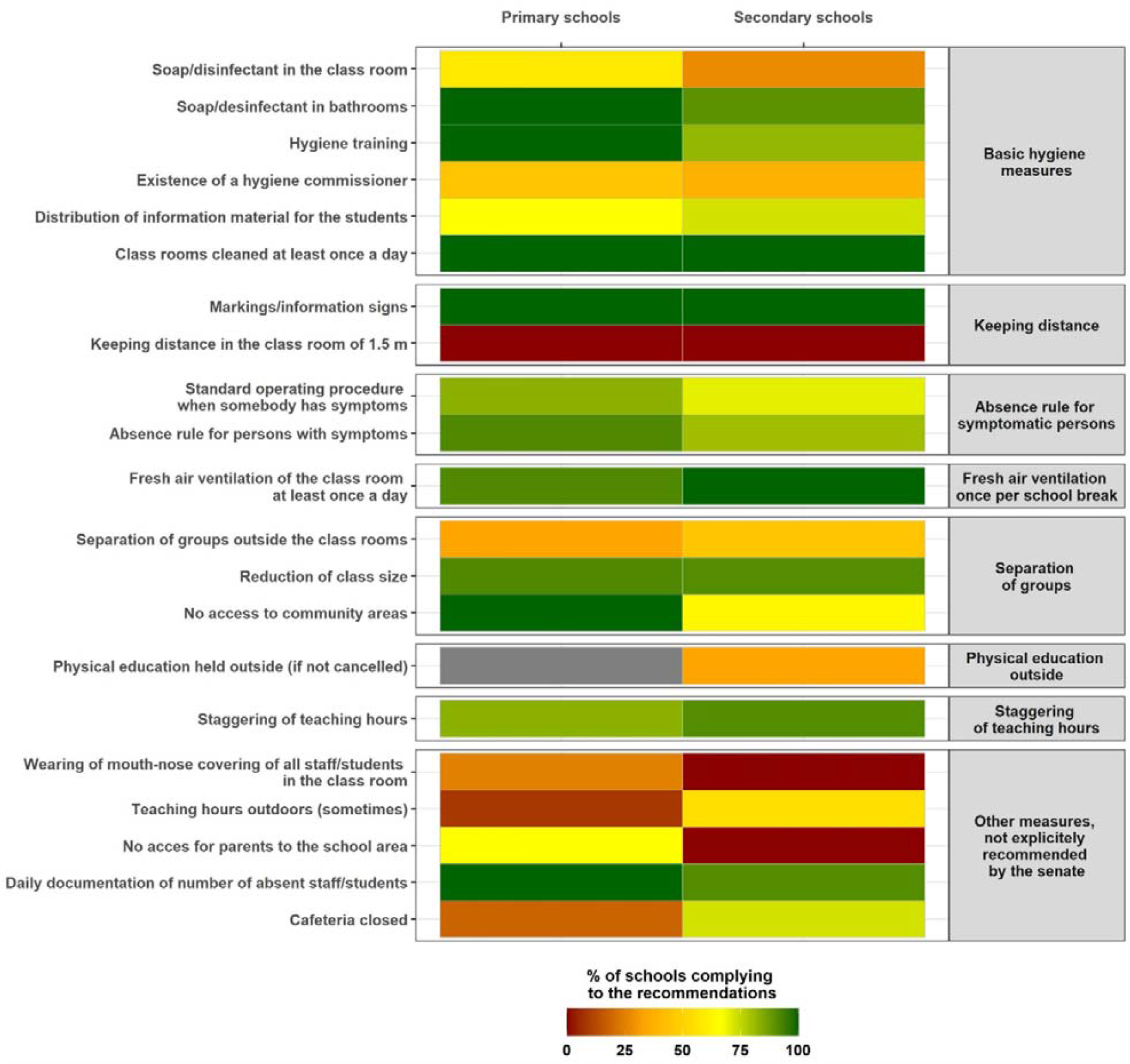
**Heat map with percentage of implementation of the preventive measures in classes, stratified by the main aspects of the senate’s recommendations valid in June 2020.**

## Discussion

In this cross-sectional study examining students and staff in Berlin schools during the early post-lockdown phase, the prevalence of SARS-CoV-2 infections (0.2%) and IgG sero-reactivity (1.3%) was low. Our study also shows that, even in summer, mild symptoms resembling a common cold are frequently reported at school (16%) and that the bulk of health impairment was not due to SARS-CoV-2 infection. Even more, the then-existing absence rule for symptomatic persons might have led to underreporting. The large proportion of affected children (even larger considering the preceding two weeks) poses a dilemma to parents, school-staff, and policy-makers. Against this backdrop, the Berlin senate issued regulations that allow regular school attendance for students with only mild and afebrile symptoms, e.g., rhinorrhoea, cough. Nevertheless, SARS-CoV-2 infection often runs an asymptomatic course in children and adolescents [25], and the mild symptoms of the one infected juvenile diagnosed in this study could have easily been overlooked. In the absence of routine testing of students, e.g., by antigen tests, asymptomatic SARS-CoV-2 infections in school presumably need to be tolerated. In this regard, low child-to-child transmission rates in educational settings are reassuring [12, 18], but it has to be seen whether this holds true at higher infection incidence. In December 2020, Germany has released antigen tests for lay-use by teachers and educators (but not by students or parents), which constitutes a good starting point considering that most individuals in school-based outbreaks so far have been adults [13]. Nevertheless, routine testing of students (taking into account asymptomatic infections), e.g., twice a week, is a desirable next step to prevent viral transmission within and out of school.

School staff reported a high prevalence of chronic conditions (44%), half of which were not specified. This may reflect the teachers’ relatively high age in the present study, like in Germany per se [26]. As a result, a substantial fraction of teachers may belong to a COVID-19-relevant risk group. A 2015 review on German teachers’ health showed that half of a teachers’ cohort was overweight and 13% obese; 48% suffered from hypertension [27], factors that may contribute to more severe COVID-19. Yet, the high reporting frequency of indeterminate chronic conditions might also point to a more general self-awareness of vulnerability among teachers. One-third of teachers reportedly experience exhaustion and high emotional workload [27], enforcing perceived vulnerability and susceptibility to health hazards. Accordingly, school staff in our study frequently reported fear of infection and self-classified as being at high risk. This perception finds reflection in high adherence to IPC measures like hand hygiene and facemask-wearing. Policymakers should take into consideration that teachers and educators, while adapting to COVID-19 related changes, need clear instructions for a safe workspace to avoid psychological distress and unnecessary school absenteeism.

Previous studies have shown unfavourable changes in social life and physical activity due to school closures [28, 29]. Our results show that much less time was spent meeting friends compared to pre-pandemic times among both students and staff. However, our instruments were not designed for in-depth assessment of the extent of social isolation and socio-affective conditions potentially arising from that, as reported elsewhere [3, 30]. Self-reported screen time increased particularly among students. Physical activity dropped among primary school pupils, but much less among older students. Data obtained during the COVID-19 pandemic in China with its very strict lockdown showed a sharp increase in screen time, in parallel with a significant decrease in physical activity [28]. While those results are transferable to the Berlin context only to a limited extent, our findings confirm this tendency especially among younger children and give rise to concern: excessive screen time in conjunction with sedentary behaviour, snacking and weight gain [29] have been associated with cardiovascular disease risk factors such as obesity, high blood pressure and insulin resistance [6]. Depending on the duration of the pandemic, there is a risk that some of the newly established behaviours may persist, requiring parents, teachers and policy-makers to promote healthy lifestyles.

Our findings show that the implementation of IPC measures in schools is feasible, as the governmental recommendations were largely implemented, with primary schools performing better than secondary schools. This might partially reflect more dismissive attitudes towards regulations, especially on social behaviour, among older students or more flexible adaption in the commonly smaller primary schools. The latest official recommendations on school operation of November 2020 strengthen fresh air ventilation and hand hygiene, and present separate advice for primary and secondary schools in form of a four-tier system to enable adaptation to the local situation [23]. Nevertheless, preventive measures at school and class level will continuously need to be adapted, mirroring and anticipating relevant epidemic developments.

Our study has several limitations. Sample size and study period pose limitations to the generalizability of our data. Voluntary participation of both, institutions and school members and low participation rates in some facilities may have caused a selection bias. Determinants of infection could not be assessed due to the detection of a single case only. On the other hand, sample collection among children and adolescents was unproblematic as reflected by the high proportion of available specimens.

Our findings suggest that educational settings and their players are largely able to adapt to IPC measures and to changing conditions. Increased screen time as well as reduced social contacts and, partially, physical activity point to the non-infectious dangers adaptations to the pandemic bring along. Needs and situational requirements of students and teachers are to be met, including such linked to fear and behaviour. This forms the prerequisites for the comprehension of and adherence to IPC measures, which in turn determine school functioning. Ongoing follow-up examinations will show whether this can be achieved. In the meantime, regular screening of students and teachers for SARS-CoV-2 in the school setting may help to reduce both infections and uncertainties, thereby ensuring the right to education.

## Data Availability

Data are available with the authors.

## BECOSS study group members

Heike Rössig, Tanja Chylla, Mandy Kollatzsch, Andreas Hetey, Omar Alhasan, Katja Püstow, Andreas Lindner, Olga Nikolai, Mia Wintel, Karen Krüger, Franziska Kindt, Annkathrin von der Haar, Jennifer Körner (Charité – Universitätsmedizin Berlin, corporate member of Freie Universität Berlin, Humboldt-Universität zu Berlin, and Berlin Institute of Health). We thank the schools, respective staff, students and their families for enabling the study.

## Authors’ contributions

FH, TK, and FPM: study design. FH, WvL, MT, IA, SL, RH, SGB, EL, AB, AS, KvB, CB, and FPM: participants enrolment and examinations. AK, MK, MG, GEM, and JR: supervision of study logistics. FB: laboratory examinations. MAM and AR: supervision of paediatric examinations. JK, NB: supervision of assessments as to behaviour and perception. TK, VK, JS, and FPM liaised with health and educational authorities. WvL and JK led the data analysis. FH, WvL, ST, JK, and FPM led the writing of the manuscript. BECOSS study group: data collection. All authors participated in drafting the article or revising it critically for intellectual content.

## References

1. Masonbrink AR, Hurley E. Advocating for Children During the COVID-19 School Closures. Pediatrics 2020. doi:10.1542/peds.2020-1440.

2. Armitage R, Nellums LB. Considering inequalities in the school closure response to COVID-19. The Lancet Global Health. 2020;8:e644. doi:10.1016/S2214-109X(20)30116-9.

3. Brooks SK, Webster RK, Smith LE, Woodland L, Wessely S, Greenberg N, Rubin GJ. The psychological impact of quarantine and how to reduce it: rapid review of the evidence. Lancet. 2020;395:912–20. doi:10.1016/S0140-6736(20)30460-8.

4. Margaritis I, Houdart S, El Ouadrhiri Y, Bigard X, Vuillemin A, Duché P. How to deal with COVID- 19 epidemic-related lockdown physical inactivity and sedentary increase in youth? Adaptation of Anses’ benchmarks. Arch Public Health. 2020;78:52. doi:10.1186/s13690-020-00432-z.

5. Bayham J, Fenichel EP. Impact of school closures for COVID-19 on the US health-care workforce and net mortality: a modelling study. The Lancet Public Health. 2020;5:e271–e278. doi:10.1016/S2468-2667(20)30082-7.

6. Nagata JM, Abdel Magid HS, Pettee Gabriel K. Screen Time for Children and Adolescents During the Coronavirus Disease 2019 Pandemic. Obesity (Silver Spring). 2020;28:1582–3. doi:10.1002/oby.22917.

7. Snape MD, Viner RM. COVID-19 in children and young people. Science. 2020;370:286–8. doi:10.1126/science.abd6165.

8. Regional Office for Health and Social Affairs Berlin (LAGeSo). COVID-19 in Berlin, distribution in the districts - complete overview (in German). https://daten.berlin.de/datensaetze/covid-19-berlin-verteilung-den-bezirken-gesamt%C3%BCbersicht. Accessed 24 Oct 2020.

9. Robert Koch Institute. Coronavirus Disease 2019 (COVID-19) - Daily Situation Report of the Robert Koch Institute. <https://www.rki.de/EN/Content/infections/epidemiology/outbreaks/COVID-19/Situationsberichte_Tab.html.

10. Merckx J, Labrecque JA, Kaufman JS. Transmission of SARS-CoV-2 by Children. Dtsch Arztebl Int. 2020;117:553–60. doi:10.3238/arztebl.2020.0553.

11. Goldstein E, Lipsitch M, Cevik M. On the effect of age on the transmission of SARS-CoV-2 in households, schools and the community. J Infect Dis. 2020.

12. Ehrhardt J, Ekinci A, Krehl H, Meincke M, Finci I, Klein J, et al. Transmission of SARS-CoV-2 in children aged 0 to 19 years in childcare facilities and schools after their reopening in May 2020, Baden-Württemberg, Germany. Euro Surveill 2020. doi:10.2807/1560-7917.ES.2020.25.36.2001587.

13. Otte im Kampe E, Lehfeld A-S, Buda S, Buchholz U, Haas W. Surveillance of COVID-19 school outbreaks, Germany, March to August 2020. Euro Surveill 2020. doi:10.2807/1560-7917.ES.2020.25.38.2001645.

14. Isphording IE, Lipfert M, Pestel N. School Re-Openings after Summer Breaks in Germany Did Not Increase SARS-CoV-2 Cases. Institute of Labor Economics. 2020;Discussion Paper No. 13790.

15. Ludvigsson JF. Systematic review of COVID-19 in children shows milder cases and a better prognosis than adults. Acta Paediatr. 2020;109:1088–95. doi:10.1111/apa.15270.

16. Dong Y, Mo X, Hu Y, Qi X, Jiang F, Jiang Z, Tong S. Epidemiology of COVID-19 Among Children in China. Pediatrics 2020. doi:10.1542/peds.2020-0702.

17. Hippich M, Holthaus L, Assfalg R, Zapardiel Gonzalo JM, Kapfelsperger H, Heigermoser M, et al. Public health antibody screening indicates a six-fold higher SARS-CoV-2 exposure rate than reported cases in children. Med (N Y) 2020. doi:10.1016/j.medj.2020.10.003.

18. Macartney K, Quinn HE, Pillsbury AJ, Koirala A, Deng L, Winkler N, et al. Transmission of SARS-CoV-2 in Australian educational settings: a prospective cohort study. Lancet Child Adolesc Health. 2020;4:807–16. doi:10.1016/S2352-4642(20)30251-0.

19. Heavey L, Casey G, Kelly C, Kelly D, McDarby G. No evidence of secondary transmission of COVID-19 from children attending school in Ireland, 2020. Euro Surveill 2020. doi:10.2807/1560-7917.ES.2020.25.21.2000903.

20. Chu DK, Akl EA, Duda S, Solo K, Yaacoub S, Schünemann HJ, et al. Physical distancing, face masks, and eye protection to prevent person-to-person transmission of SARS-CoV-2 and COVID- 19: a systematic review and meta-analysis. The Lancet. 2020;395:1973–87. doi:10.1016/S0140-6736(20)31142-9.

21. Schwarzer R. Modeling Health Behavior Change: How to Predict and Modify the Adoption and Maintenance of Health Behaviors. Applied Psychology. 2008;57:1–29. doi:10.1111/j.1464-0597.2007.00325.x.

22. Wittmann N, Sallmon S, Meinlschmidt G. Gesundheits- und Sozialstrukturatlas für die Bundesrepublik Deutschland [Publication in German]. 2015. <https://www.berlin.de/sen/gesundheit/service/gesundheitsberichterstattung/veroeffentlichungen/spezialberichte/.

23. Berlin Senate Administration for Education, Youth and Family. Musterhygieneplan Corona für die Berliner Schulen [Publication in German]. 2020. <https://www.berlin.de/sen/bjf/coronavirus/aktuelles/schrittweise-schuloeffnung/20200623_musterhygieneplan-corona-fuer-die-berliner-schulen.pdf.

24. Harris PA, Taylor R, Minor BL, Elliott V, Fernandez M, O’Neal L, et al. The REDCap consortium: Building an international community of software platform partners. J Biomed Inform. 2019;95:103208. doi:10.1016/j.jbi.2019.103208.

25. Castagnoli R, Votto M, Licari A, Brambilla I, Bruno R, Perlini S, et al. Severe Acute Respiratory Syndrome Coronavirus 2 (SARS-CoV-2) Infection in Children and Adolescents: A Systematic Review. JAMA Pediatr. 2020;174:882–9. doi:10.1001/jamapediatrics.2020.1467.

26. OECD. Bildung auf einen Blick 2020: OECD-Indikatoren. 2020. https://www.oecd-ilibrary.org/education/bildung-auf-einen-blick-2020-oecd-indikatoren_6001821nw;jsessionid=IuMPVy18f62Xhu7lTSzxz-Qj.ip-10-240-5-151.

27. Scheuch K, Haufe E, Seibt R. Teachers’ Health. Dtsch Arztebl Int. 2015;112:347–56. doi:10.3238/arztebl.2015.0347.

28. Xiang M, Zhang Z, Kuwahara K. Impact of COVID-19 pandemic on children and adolescents’ lifestyle behavior larger than expected. Prog Cardiovasc Dis. 2020;63:531–2. doi:10.1016/j.pcad.2020.04.013.

29. Ammar A, Brach M, Trabelsi K, Chtourou H, Boukhris O, Masmoudi L, et al. Effects of COVID-19 Home Confinement on Eating Behaviour and Physical Activity: Results of the ECLB-COVID19 International Online Survey. Nutrients 2020. doi:10.3390/nu12061583.

30. López-Bueno R, López-Sánchez GF, Casajús JA, Calatayud J, Tully MA, Smith L. Potential health-related behaviors for pre-school and school-aged children during COVID-19 lockdown: A narrative review. Prev Med. 2020:106349. doi:10.1016/j.ypmed.2020.106349.

